# Performance of Gazelle COVID-19 point-of-care test for detection of nucleocapsid antigen from SARS-CoV-2

**DOI:** 10.1101/2022.03.23.22272094

**Authors:** Amrish Mehta, Bri Spencer, Ashok Garg, Pradeep Kalmorge, Raúl A. Ocasio Gonzalez, Kate Taussig, Ryan R. Fortna

## Abstract

SARS-CoV-2 antigen assays offer simplicity and rapidity in diagnosing COVID-19. We assessed the clinical performance of Gazelle COVID-19 test, a fluorescent lateral flow immunoassay with an accompanying Reader utilizing image-recognition software for detection of nucleocapsid antigen from SARS-CoV-2. We performed a prospective, operator-blinded, observational study at 2 point-of-care (POC) sites. Nasal swab specimens from symptomatic patients were tested with Gazelle COVID-19 test and real-time polymerase chain reaction (RT-P CR) assay. Overall, data from 1524 subjects was analyzed, and 133 were positive by RT-PCR. Mean (range) age of participants was 34.7 (2-94) years and 570 (37.4%) were female. The sensitivity and the specificity of the Gazelle COVID-19 test were 96.3% and 99.7%. The PPV of Gazelle COVID-19 test was 97.0%, NPV 99.6%, and accuracy 99.4%. In POC settings, Gazelle COVID-19 test had high diagnostic accuracy for detection of SARS-CoV-2 in nasal swab samples of symptomatic subjects suspected of COVID-19.

## INTRODUCTION

The World Health Organization (WHO) declared coronavirus disease 2019 (COVID-19), the disease caused by severe acute respiratory syndrome coronavirus 2 (SARS-CoV-2), a pandemic health emergency [1]. The high rate of community and institutional transmission [2] of SARS-CoV-2 infection underscores the need for rapid and accurate detection of active infection. In a survey of unmet needs for COVID-19 tests in health and social care settings, hospitals identified COVID-19 testing as the second highest unmet need, with the greatest priority being a test for symptomatic patients presenting to hospitals for infection control [3]. Studies have shown that of the various steps required in the process of testing and diagnosing an active infection, minimizing testing delay had the largest impact on reducing onward transmissions [4, 5]. Current diagnostic tests for the SARS-CoV-2 detect either nucleic acid or antibody of viral protein [6]. Several molecular assays based on reverse transcriptase polymerase chain reaction (RT-PCR) have been developed to detect and quantify SARS-CoV-2 RNA. However, COVID-19 patients can continue to shed viral RNA well beyond clinical recovery and positive RT-PCR does not necessarily indicate infectiousness [7]; RT-PCR testing detects residual genome, and a test of actual infectivity (viable virus) is needed. Furthermore, RT-PCR is expensive, has a relative long turn-around-time, require trained personnel, and is often available only in laboratory set-ups that provide centralized services. In February 2020, WHO identified rapid testing with point-of-care (POC) diagnostics as a number one priority to address the COVID-19 pandemic [8] and the US Food and Drug Administration (FDA) permitted licensed laboratories to report in-house developed SARS-CoV-2 diagnostic tests [9].

The Gazelle COVID-19 Test is a fluorescent lateral flow immunoassay (FIA) for the qualitative detection of nucleocapsid antigen from SARS-CoV-2. The Gazelle platform includes a Reader which is capable of reading both colorimetric and certain fluorescent lateral flow assays and is adapted from the commercially available hemoglobin variant reader, using derived image-recognition software [10]. Here, we present the results from an observational study to assess the diagnostic performance of Gazelle Covid-19 Test, compared to RT-PCR.

## METHODS

### Study Design and Population

We conducted a prospective, observational study to determine the diagnostic performance of Gazelle Covid-19 Test, compared to RT-PCR. The study was conducted in two different regions from 23 August 2021 through 12 November 2021. The first site was at Northwest Laboratory’s COVID-19 drive-through screening site located at the Bellingham Airport, Washington, USA and the second one was at Bio Diagnostics laboratory, Pune and Acu-MDx Laboratory and Research Center Pvt. Ltd Diagnostics Lab, Mumbai, India. The objective of the study was to assess Gazelle COVID-19 Test performance (positive percent agreement [PPA] and negative percent agreement [NPA]) in comparison to RT-PCR using dual mid-turbinate nasal swab samples from symptomatic subjects (within five days of onset of symptoms) at point of care.

Participants were recruited, screened, and enrolled by the Investigator’s team at each site as per the inclusion and exclusion criteria pre-specified in the study protocol. All consecutive patients who visited the facilities for a Covid test and who met inclusion criteria were enrolled in the study and informed consent was obtained. Operators of the Gazelle COVID-19 Test and the RT-PCR test were blinded to the results on the different test platform. Additionally, operators tested a set of blinded, contrived samples to ensure consistency of performance between operators at the Bellingham location

All methods were performed in accordance with the relevant guidelines and regulations. This paper was drafted according to STROBE and STARD guidelines to ensure the quality of reporting [11, 12].

### Sample Collection

Direct human nasal swab samples were collected from symptomatic subjects and tested immediately on Gazelle at each site. The swab for RT-PCR was collected immediately before the swab for the Gazelle test. The swabs for RT-PCR were stored at ambient temperature in Brooks tubes [should probably provide the item number here] with 0.9% sterile saline according to Northwest Laboratories protocol, then transported to the laboratory at the end of the day for testing within 24 hours of collection. The swabs collected in India were also stored in sterile saline and tested within 24 hours of collection.

### Gazelle Covid-19 Test

Gazelle COVID-19 (Figure 1) is a fluorescent lateral flow immunoassay and accompanying Reader intended for the detection of nucleocapsid antigen from SARS-CoV-2 in nasal swab specimens who are suspected of COVID-19 by their healthcare provider within 5 days of symptom onset. The Gazelle COVID-19 Test detects COVID-19 nucleocapsid protein in dual nasal mid-turbinate swab specimens. The Reader is equipped with LEDs and is capable of reading both colorimetric and certain fluorescent lateral flow assays.

**Figure 1.**
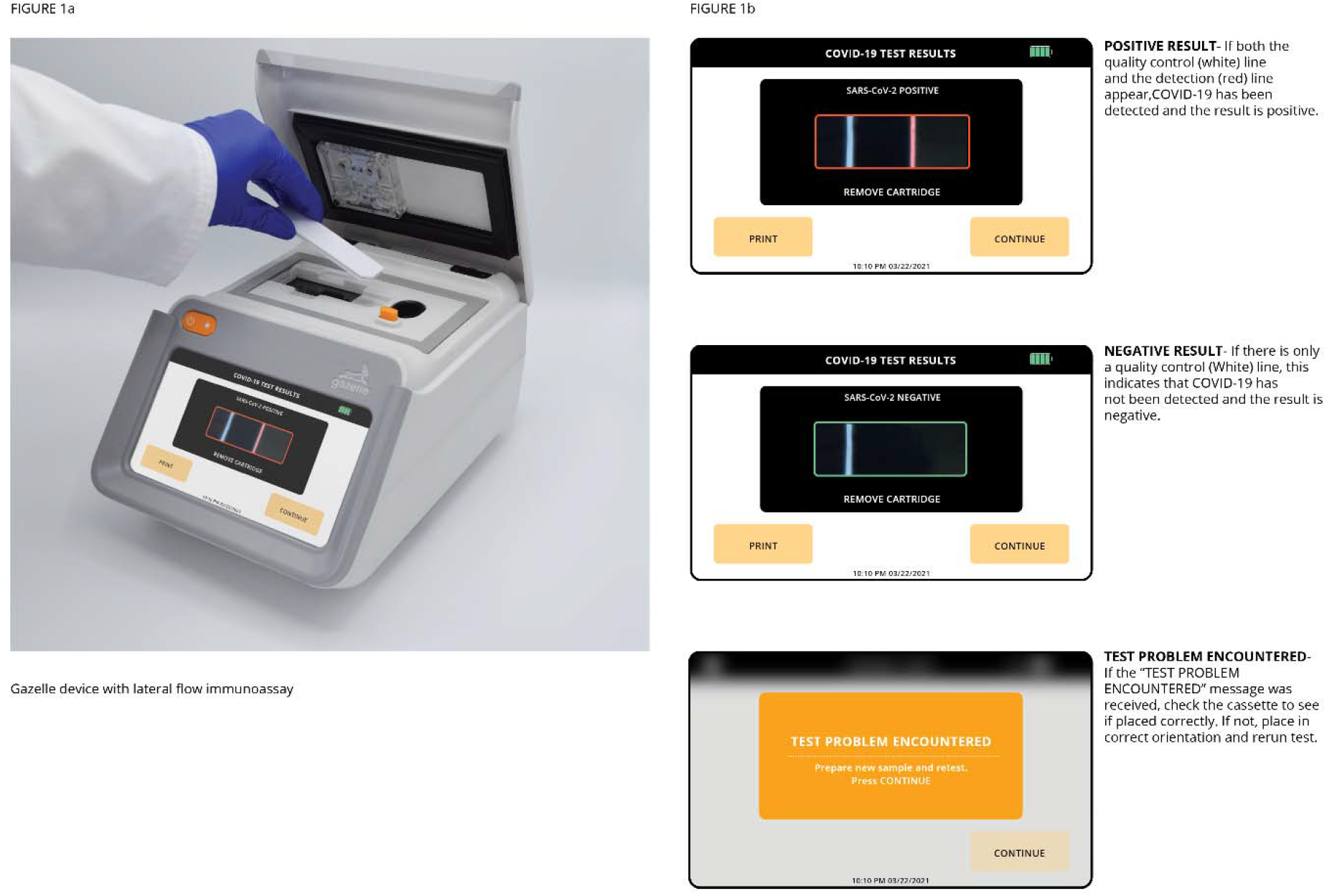
Gazelle Covid-19 Reader and interpretation of results

### RT-PCR Procedure

#### Northwest Laboratories, USA

Confirmation of COVID-19 status was done by using the TaqPath™ COVID-19 Combo Kit (ThermoFisher Scientific, Pleasanton, CA, USA - Document: MAN0019181 Rev. A.0. March 12, 2020.). RT-PCR assays and analysis were performed following the instructions of the manufacturer using Northwest Laboratory’s 7500 Fast Real-Time PCR System Thermocycler (Applied Biosystems, CA, USA). This system was validated via correlation studies with the Washington State Public Health Laboratory using a test system authorized for emergency use by the FDA. Briefly, the conditions of the thermal profile followed several specific steps: (1) 25°C for 2 min; (2) 53°C for 10 min; (3) 95°C for 2 min (4) 40 cycles at 95°C for 3 s, and 60°C for 30 s. According to the instructions of the manufacturer in N-target, the Ct value <37 or two replicates with Ct values of 38 were considered positive; Ct values >39 were considered negative. An exogenous control was included in all reactions using primers and probe specifically directed at MS2-Phage genomic RNA, obtained from TaqPath™ COVID-19 Combo Kit. Furthermore, positive, negative, and non-template controls were included in each run. A Ct value <30 in the positive control and no Ct values in negative and non-template controls were considered to validate runs.

### Bio Diagnostics laboratory and Acu-MDx Laboratory and Research Center Pvt. Ltd Diagnostics, India

Confirmation of COVID-19 status was done by using the Taq Path Covid RT-PCR kit (Applied Biosystems; Document No: 100033995 Rev. B.0.Pub No. MAN0014069). RT-PCR assays and analysis were performed following the instructions of the manufacturer using Quantstudio 5 RT-PCR Instrument (Applied Biosystems). Briefly, the conditions of the thermal profile followed several specific steps: (1) 25°C for 2 min; (2) 53°C for 10 min; (3) 95°C for 2 min (4) 40 cycles at 95°C for 3 s, and 60°C for 30 s. According to the instructions of the manufacturer in N-target, the Ct value <35; Ct values >35 were considered negative.

### Data collection and Statistical Analysis

Operators of the Gazelle test were the employees certified by the respective laboratories at taking nasal swabs with no formal training. The operators were provided the Instructions for Use (IFU). No additional training on the Gazelle test was provided to the operators. Following Gazelle COVID-19 Test, data from the Gazelle Reader was sent to Hemex to be analyzed by the Gazelle Covid-19 algorithm. RT-PCR results were recorded at the testing laboratory according to its procedures. They were also recorded in a Google spreadsheet provided by the sponsor. Other Case Report Form information was entered by the test site and then copied into the Google spreadsheet.

All test results recorded in the spreadsheets were reviewed and verified for completeness and entry accuracy by the Investigator and Hemex Monitor (Hemex employee responsible for monitoring study quality). Test results were verified daily to ensure they were complete and error free.

The diagnostic performance Gazelle Covid-19 test was assessed using sensitivity, specificity, positive predictive value (PPV) and negative predictive value (NPV). Data was analyzed for sensitivity or PPA (i.e., the probability that the assay will be positive when the comparator assay is positive; sensitivity = comparator positive (TP)/(TP + false negative [FN]) ×100%) and specificity or NPA (i.e., the probability that the assay will be negative when the comparator assay is negative; specificity = comparator negative (TN)/(TN + (false positive [FP]) ×100%).

## RESULTS

A total of 1090 potential participants were screened in India and 458 were screened at Bellingham site in the United States. Patient screening and enrolment details are presented in the flow chart (Figure 2).

**Figure 2.**
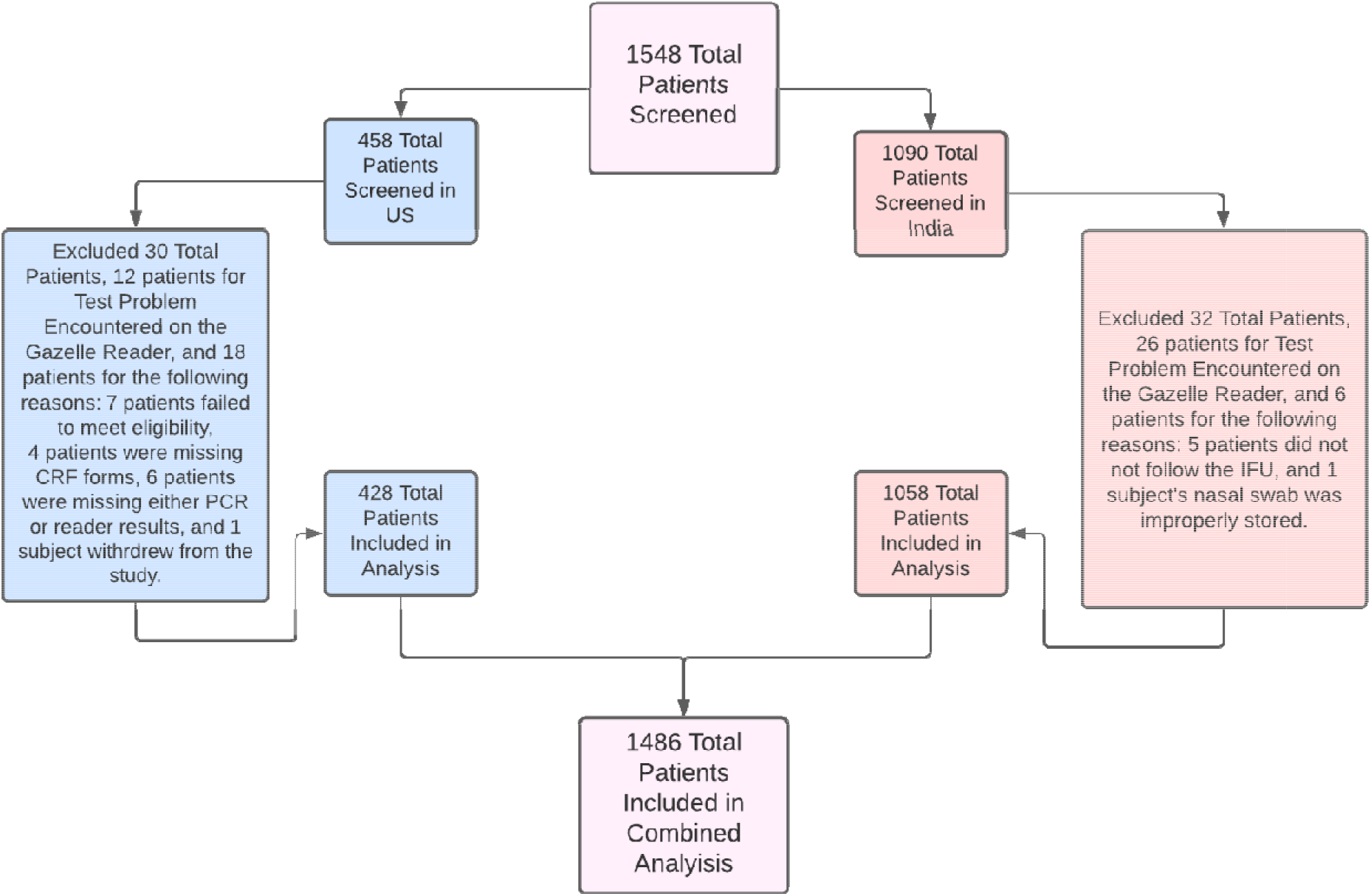
Patient screening and enrolment flowchart

Overall, data from 1486 participants was analyzed. Mean (range) age of study participants from the Bellingham site was 32.8 (2-90) years and 250 (58.4%) were female. Mean (range) age of study participants from India was 35.5 (4-94) years and 304 (28.7%) were female. A total of 133 tests were RT-PCR positive and 1353 were RT-PCR negative. Gazelle Test results and performance in comparison to RT-PCR is presented in **Table 1 and Table 2**, respectively.

**Table 1.** Results of Gazelle Covid-19 Test in comparison to RT-PCR

| **RT-PCR confirmed COVID-19** |  |  |  |  |
| --- | --- | --- | --- | --- |
| **United States** |  |  |  |  |
|  |  | Positive | Negative | Total |
| Gazelle Covid-19 Test | Positive | 44 | 1 | 45 |
|  | Negative | 2 | 381 | 383 |
|  | Total | 46 | 382 | 428 |
| **India** |  |  |  |  |
|  |  | Positive | Negative | Total |
| Gazelle Covid-19 Test | Positive | 85 | 3 | 88 |
|  | Negative | 3 | 967 | 970 |
|  | Total | 88 | 970 | 1058 |
| **Combined** |  |  |  |  |
|  |  | Positive | Negative | Total |
| Gazelle Covid-19 Test | Positive | 129 | 4 | 133 |
|  | Negative | 5 | 1348 | 1353 |
|  | Total | 134 | 1352 | 1486 |

**Table 2.** Performance of Gazelle Covid-19 Test in comparison to RT-PCR

|  | **United States** | **India** | **Combined % (n/N)** |
| --- | --- | --- | --- |
| Sensitivity | 95.7% (44/45) | 96.6% (85/88) | 96.3% (129/134) |
| Specificity | 99.7% (381/383) | 99.7% (967/970) | 99.7% (1348/1352) |
| Positive Predictive Value (PPV) | 99.3% (425/428) | 99.7% (967/970) | 97.0% (129/133) |
| Negative Predictive Value (NPV) | 97.8% (44/45) | 96.6% (85/88) | 99.6% (1348/1353) |
| Accuracy | 99.5% (381/383) | 99.7% (967/970) | 99.4% (1477/1486) |

A total of 23 operators performed 428 tests (range, 1-59) at the Bellingham site. As a quality control measure, consistency of performance among test operators was also assessed at the Bellingham site. A total of 24 Gazelle tests (4 Gazelle tests per sample) were run across 6 contrived samples (A, B, C D, E, and F). Each of the 4 operators assessed ran one Gazelle test on each of the 6 contrived samples. Three of the contrived samples were negative (samples A, D, and F), and 3 were <2x limit of detection positive (samples B, C, and E). The concentration of the positive samples was 10-50% tissue culture infective dose/mL. The tests were run interspersed with the clinical samples across 2 days (20 October 2021 and 21 October 2021). Gazelle Covid-19 test showed high inter-operator test operator agreement. Gazelle also showed 100% accuracy on all the contrived sample testing.

## DISCUSSION

Rapid-result testing for SARS-CoV-2 has value in many clinical settings that may not have access to specialized laboratory equipment or personnel and have a need for immediate decision-making or counseling the patient according to the results obtained. Here we describe the performance of Gazelle Covid-19 test for the qualitative detection of SARS-CoV-2 in nasal swab samples from symptomatic subjects. Gazelle Covid-19 test demonstrated high diagnostic potential with low false positive/negative rates and a detection accuracy of 99.4% compared to the RT-PCR. Gazelle Covid-19 test meets the criteria recommended in WHO interim guidance for diagnosis of SARS-CoV-2 infection (at least 80% sensitivity and 97% specificity) with rapid antigen assays [13].

An optimal Covid-19 test would detect replicating or transmissible virus, avoiding prolonged positivity of RT-PCR after infectiousness has resolved. There is currently no gold standard test that identifies infectious subjects with replicating or transmissible virus. SARS-CoV-2 nucleocapsid antigen is generally detectable in nasal swabs during the acute phase of infection [14]. Positive results indicate the presence of viral antigens, but clinical correlation with patient history and other diagnostic information is necessary to determine disease status. In this context, antigen testing has been shown to have higher accuracy among specimens with positive viral culture, correlating with the period when infectious virus is present [15].

Persons who know their positive test result can isolate sooner, and where appropriate contact tracing can be initiated sooner and be more effective. Asymptomatic SARS-CoV-2 infected subjects were not included in the study, and lower viral loads are likely to reduce sensitivity relative to PCR, though these cases are also less likely to transmit (REF). A study to assess the performance of Gazelle Covid-19 test in an asymptomatic population is currently underway. This study adds to the growing evidence of the performance of different rapid SARS-CoV-2 antigen detections, especially in the community settings. Gazelle Covid-19 test is easy to perform with minimal training or previous laboratory testing experience and is thus a feasible solution to implement at sites requiring a POC solution. Inter-operator test operator agreement in the target clinical population and environment indicated that the device operation and the results are reproducible.

This study has some limitations. subjects with invalid results were not able to be retested because participants left the facility before test results were available. A total of 2.6% of the initial Gazelle Covid-19 test results were either invalid or canceled and would have required patient retesting in accordance with the IFU. Re-collection of samples and retesting would be expected to further reduce the invalid rate, but this was not possible during our study. Discrepant results observed between the Gazelle Covid-19 test and reference RT-PCR could not be further clarified, as further samples were not available for retesting.

In summary, herein we have shown that the Gazelle Covid-19 test using a nasal swab collection method is highly accurate and has close concordance with RT-PCR based assay for detection of SARS-CoV-2 in a POC setting. Gazelle Covid-19 test has potential to quickly and effectively screen all symptomatic patients suspected of COVID-19 at POC, enabling rapid detection and isolation of people most likely to pose an infectious risk.

## Data Availability

The datasets generated during and/or analyzed during the current study are available from the corresponding author on reasonable request.

## ETHICS STATEMENT

Institutional Review Board approval was obtained by the sponsor before starting the study in the United States (WCG IRB Tracking number: 20212857). Study protocol from India was approved by the Ethics Committee of International Institute of Sleep Sciences (Reg No: ECR/177/Indt/MH-2014/RR-19). Informed consents were obtained from all study participants.

## FUNDING

The funding for the study was provided by Hemex Health, Inc., USA.

## ACKNOWLEDGMENTS

None

## CONFLICT OF INTEREST

ROG and KT were hired as consultants by Hemex Health to execute this study. AM is a board member of HemexDx, a subsidiary of Hemex Health in India.

## AUTHOR CONTRIBUTIONS

AM, AG and PK helped with the planning and execution of clinical study in India and BS, ROG, KT and RRF helped with the planning and execution of the clinical study in the US. AG, PK, BS, ROG and KT helped with subject recruitment, nasal swab collection, and testing. All the authors authored and edited the manuscript and helped in preparing the tables, figures, figure captions, and supplementary information.

## Notes

### Clinical Trial

NCT04987918

